# NTDscope: A multi-contrast portable microscope for disease diagnosis

**DOI:** 10.1101/2025.03.17.25324136

**Authors:** María Díaz de León Derby, Zaina L. Moussa, Carlos F. Ng, Abdul M. Bhuiya, Joana P. Cabrera, Dipayan Banik, Charles B. Delahunt, Matthew D. Keller, Anne-Laure M. Le Ny, Jaime Garcia-Villena, Elena Dacal, David Bermejo-Peláez, Daniel Cuadrado, Miguel Luengo-Oroz, Hugues C. Nana Djeunga, Joseph Kamgno, Linda Djune Yemeli, Victor Pahl, Saskia D. Davi, Rella Zoleko Manego, Michael Ramharter, Isaac I. Bogoch, Jean T. Coulibaly, Amena Khatun, Mamun Kabir, Zannatun Noor, Rashidul Haque, Neil A. Switz, Daniel H. Friedman, Michael V. D’Ambrosio, Daniel A. Fletcher

## Abstract

Accurate diagnostics are essential for disease control and elimination efforts. However, access to diagnostics for neglected tropical diseases (NTDs) is hindered by limited healthcare infrastructure in many NTD-endemic regions, as well as by reliance on time- and labor-intensive diagnostic methods, such as smear microscopy. New diagnostic tools that are portable, rapid, low-cost, and meet World Health Organization (WHO) sensitivity and specificity targets are urgently needed to accelerate NTD control and elimination programs. Here, we introduce the NTDscope, a portable microscopy platform that enables point-of-care imaging and automated detection of parasites and other pathogens in patient samples. The NTDscope builds on and extends the capabilities of the LoaScope, a device that turned the camera of a mobile phone into a microscope and used on-board image processing to automatically quantify *Loa loa* microfilariae burden in whole blood samples. The NTDscope replaces the mobile phone of the LoaScope with a system-on-module (SOM) that enables the integration of multiple imaging modalities in a single package designed to improve robustness and expand applications. In this work, we demonstrate use of the NTDscope as a portable brightfield, darkfield, and fluorescence microscope for samples including microfilariae and helminth eggs. We also show that the device can be used to quantify molecular assays, such as a lateral flow test and a CRISPR-Cas13a-based assay. The ability to combine diagnostic capabilities of conventional microscopy with molecular assays and machine learning in a single device could expand access to diagnostics for populations in NTD-endemic areas and beyond.

**Author summary:** Neglected tropical diseases (NTDs) impact one billion of the world’s most vulnerable individuals. Diagnostics are a necessary part of NTD disease control and elimination efforts, but identifying infected individuals remains a challenge. Here we present the NTDscope, a portable multi-contrast microscope designed to diagnose multiple NTDs at the point-of-care. We show that the NTDscope can be used to detect the parasitic worm *Loa loa* in videos of whole blood samples and parasitic eggs in images of urine and stool samples. The NTDscope can also be used to image thick blood smears in disposable capillaries and serves as a lateral flow assay reader. In addition to brightfield and darkfield imaging, fluorescence imaging on the device enables molecular assays based on CRISPR-Cas enzymes. This portable (<1 kg), field-friendly device—tested in Cameroon, Gabon, Côte d’Ivoire, and Bangladesh—has the potential to become a platform technology that addresses diagnostic needs for multiple NTDs and could serve as a key element of decentralized healthcare in the future.

## Introduction

Despite ongoing disease control and elimination efforts, neglected tropical diseases (NTDs) impact over one billion of the world’s most vulnerable individuals. For some NTDs, identification of individuals in need of treatment or verification of mass drug administration efficacy remains a major challenge [1–3]. Limited healthcare infrastructure in some NTD-endemic regions means that diagnostics must be brought to individuals, rather than individuals going to local clinics for testing. In the absence of point-of-care diagnostics, samples must be transported to a centralized facility for preparation and analysis, introducing long delays and challenges returning test results to specific individuals. While lateral flow assays and other molecular tests have simplified identification of some diseases, point-of-care molecular tests for NTDs including schistosomiasis, soil-transmitted helminths, and filarial infections are in limited use or still in development [4–12].

As a result, diagnosis of many NTDs continues to rely on conventional methods, such as thick smear microscopy for filarial parasites like *Loa loa* and urine microscopy for *Schistosoma haematobium* eggs. New approaches that are faster, cheaper, and more accurate than conventional microscopy are urgently needed to accelerate NTD control and elimination programs, with an emphasis on diagnostics that can enable rapid point-of-care testing of individuals, support starting and stopping decisions for mass drug administration (MDA) campaigns, and monitor re-emergence and spread of infections [1–3, 13–17].

Diagnostic strategies that can detect multiple NTDs are particularly compelling, given the limited resources in many NTD-endemic regions [14, 17, 18] and the need to keep costs per test as low as possible. It may be more cost-effective to have a single device diagnose multiple diseases than to develop separate assays using separate equipment. For example, automated slide-scanning microscopes that can image different types of prepared patient samples (e.g. blood, urine, or stool) have recently been developed [19–24], offering versatility in pathological diagnostics. However, these devices are often bulky, sometimes requiring the use of a separate computer, and are constrained by sample preparation methods that are typically slow and labor intensive, such as fixing and staining a blood smear. They also require technical expertise to run and interpret data, though machine learning algorithms are being developed to automate interpretation [19–21, 23, 25–34].

We previously developed a point-of-care diagnostic device called the LoaScope to identify patients who should not be treated during onchocerciasis MDA campaigns to avoid the potential for serious adverse events that are known to occur at high *Loa loa* microfilarial loads [35]. The LoaScope was designed to be a portable, field-friendly microscope that involved minimal sample preparation and provided rapid (<5 min) point-of-care quantification of microfilaria in peripheral whole blood [36]. The device was successfully used to quantify *Loa loa* microfilarial load of >16,000 people in Cameroon as part of a Test-and-Not-Treat program to resume ivermectin MDA for onchocerciasis in Loa-endemic regions [37]. The LoaScope harnessed the camera of a mobile phone and a reversed lens optical system [38], together with 3D-printed components and consumer electronics for illumination and sample translation. Sample preparation and imaging were simplified by loading patient peripheral blood into a disposable capillary with a rectangular cross-section that could be directly inserted into the LoaScope for imaging [36].

Since the LoaScope has sufficient resolution to capture images of parasite eggs, we tested whether the device could be used for detection of *Schistosoma haematobium* eggs in urine samples. We developed a tapered version of the capillary that captures particles greater than 20µm and used it to filter and concentrate *S. haematobium* eggs from patient samples [32]. We used a new version of the LoaScope with darkfield illumination capabilities, called the SchistoScope, to image the concentrated eggs in Ghana and Côte d’Ivoire [32, 39, 40]. Darkfield images proved to be useful for identification of eggs, demonstrating that the LoaScope could expand its applications to other NTDs [34].

Here, we present the next generation of the LoaScope, renamed the NTDscope for its additional capabilities that expand the set of potential diagnostic applications. The NTDscope provides multi-contrast portable imaging for both microscopy applications and molecular assay detection. Rather than using a phone itself, the NTDscope uses specific mobile phone components, such as camera modules, a rechargeable battery, and sensors, allowing the parts to be configured into an integrated and portable platform. As in the LoaScope, the new device takes advantage of a low-cost reversed lens optical system with a wide field of view (FOV) [38] and uses single-axis motion for acquisition of multiple images and videos of samples in capillaries. User-facing software is based on Android, and cloud connectivity enables data upload and remote software updates. Upgraded computing capabilities of the NTDscope support the use of machine learning (ML) algorithms for automated disease identification.

Below, we outline the technical specifications of the NTDscope and demonstrate both current and future use cases, including video capture of live microfilarial and *Schistosoma* samples, multi-contrast imaging of parasitic eggs, imaging of stained samples, and detection of molecular assays. The NTDscope has the potential to become a platform technology that addresses image-based and molecular diagnostic needs for multiple NTDs and serves as a key element of decentralized healthcare in the future.

## Materials and methods

### Ethics statement

This work contains images and videos of patient samples from four separate studies conducted in Cameroon, Gabon, Côte d’Ivoire, and Bangladesh.

The study in Cameroon was conducted in the Awae Health District, located in the Mefouet-Afamba Division, between September and October of 2023. Finger prick blood was collected from study participants. Ethical permission for this study was granted by the Centre Regional Ethical Committee for Research on Human Health (CRERSH-Ce; CE N°0094/CRERSHC/2023) and administrative authorization was obtained from the Centre Regional Delegation for Public Health of the Ministry of Public Health. The study participants signed informed consents.

The study in Gabon was conducted in the Sindara and Lambaréné regions in August of 2023. Finger prick blood was collected from study participants. Ethical permission for this study was granted by the Comité d’ethique Institutionnel du CERMEL (CEI-026/2022). Adults 18 years or older were invited to participate and provided a written informed consent.

The study in Côte d’Ivoire was conducted around the town of Azaguié in January of 2024. Ethical permission was granted by the University Health Network, Toronto, Canada (REB #21-5582) and the Comité National d’Éthique des Sciences de la Vie et de la Santé, Abidjan, Côte d’Ivoire (REB #186-21/MSHPCMU/CNESVS-km). Ethical permission was also granted by the local health district officer. Community members aged five and older were asked to provide a urine sample. Adults provided written consent and assenting children who had written consent from a parent or guardian were included.

The study in Bangladesh was conducted from October 2019 - December 2020 in the Rohingya Refugees camp in Bangladesh. Ethical permission for this study was granted by the International Centre for Diarrhoeal Disease Research, Bangladesh (icddr,b) Institutional Review Board (research protocol #PR-19014). Children who were residents of the camp and seeking care from icddr,b-operated Diarrhoea Treatment Centre were asked to participate by providing stool samples. Children were included if they assented and had written consent from a parent or guardian. Banked stool samples were used for the purposes of this paper; participants and their guardians were informed that unused samples would be stored for future use, and at any time participants and their guardians could contact the study team if they wished for the samples to be discarded and not saved.

### External Dimensions

The NTDScope has dimensions of 200mm (H) x 110mm (W) x 60mm (D) and weighs ∼880 grams. A diagram of the device and its main components is found in Fig 1. The injection-molded top housing holds a rechargeable Lithium-ion 3.7V battery and an LCD screen display. The bottom housing is composed of two pieces: a structural carriage and an outer protective housing with an over-molded elastomeric shell. The bottom housing holds the electrical boards, optical system, and a heat sink for the System-On-Module (SOM) and illumination board. Charging of the battery is done via USB-C, and battery life in field settings in Cameroon and Gabon has been observed to be around 6 hours.

**Fig 1.**
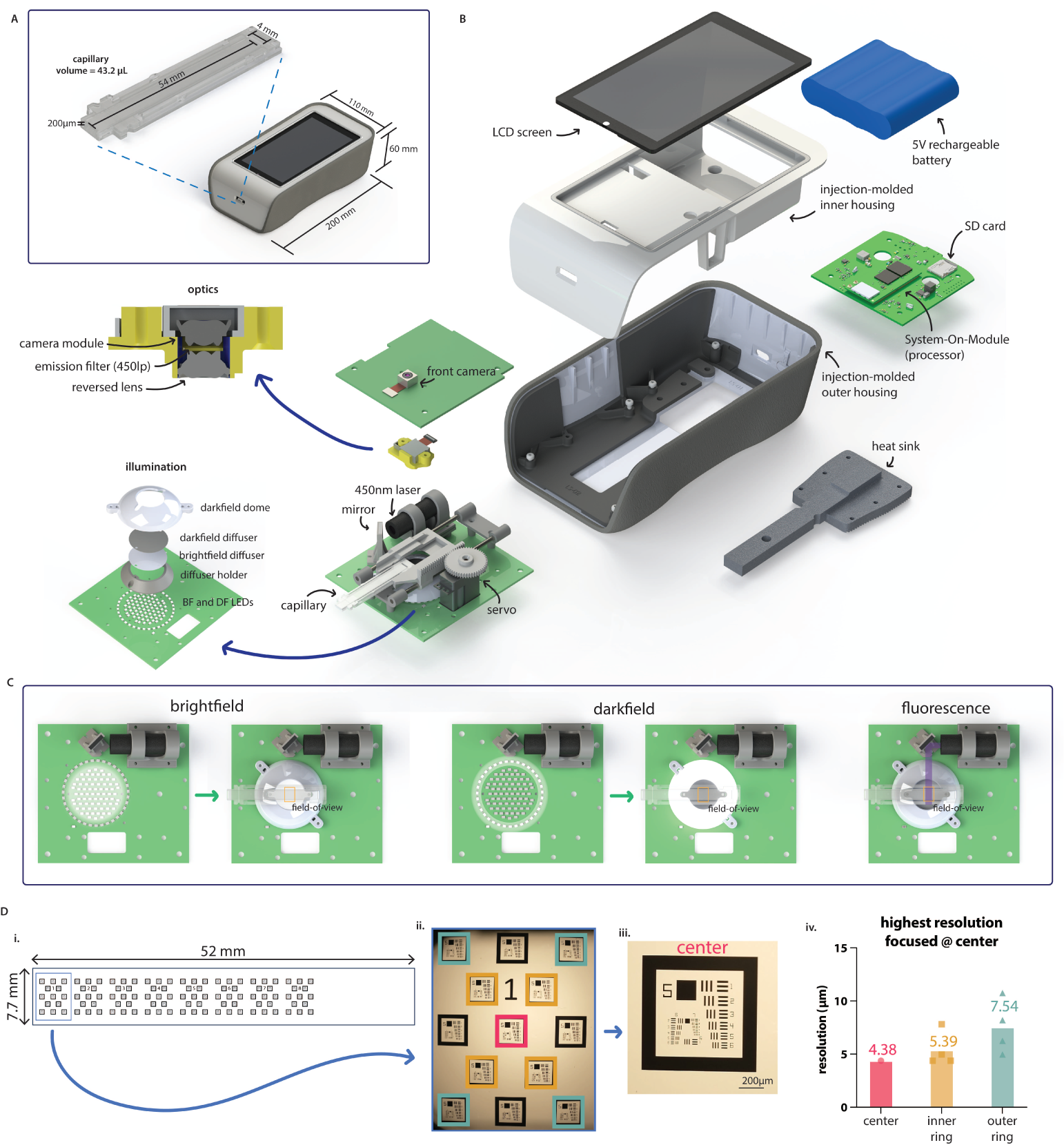
NTDscope design, illumination and resolution. **A**: Diagram showing NTDscope and straight channel capillary external dimensions. **B**: Exploded view of NTDscope and main components. **C**: NTDscope illumination modes, showing brightfield, darkfield and fluorescence illumination. **D**: NTDscope resolution measurements, showing design of custom slide with USAF resolution targets (D.i.), image of USAF resolution target array acquired on NTDscope used to measure resolution, with the resolution targets used in calculations color coded with pink (center), yellow (inner ring) and green (outer ring) (D.ii.), close up of center resolution target (D.iii.), and resolution measurements at each target location (D.iv.). Data was processed and visualized using GraphPad Prism 10.

### Electronics and Processor

Instead of a mobile phone, the NTDscope uses a series of three electronics boards containing connections to power, processors, sensors, and actuators. It is based around a System-On-Module (SOM)(Open-Q 660µSOM, Intrinsyc) that features a Qualcomm SDM660 processor. The main control carrier board (CCB) connects the battery, LCD screen, SD card for storage (256GB), and the SOM board. The illumination board has connections for the brightfield and darkfield LEDs and laser for fluorescence detection, as well as to a servomotor used to move the capillary carriage. Certain hardware components, including the carriage translation system, the fluorescence illumination components, and the servomotor, are also attached to the illumination board. Finally, the camera board contains connections for the sample and front cameras.

### Imaging System

The system uses a SONY IMX586 sensor, with a 6.4mm x 4.8mm active area comprised of 48 megapixels of 0.8µm pitch, arranged in a quad-Bayer array which thus has an effective pixel pitch of 3.2µm for the red and blue pixels and 2.3µm for the (45° rotated compared to red and blue) green pixel array [41]. The optical train is a reversed-lens configuration using Largan Precision lenses and provides a measured on-axis resolution of ≲4.4µm (Fig 1D) with a (measured) magnification of M=1.00. The optical image area exceeds the sensor dimensions, resulting in a sensor-limited field of view. The system also includes an external front camera, with a SONY IMX214 sensor (13 megapixels and 1.12µm effective pixel size [42]), which is not a part of the reversed lens system but can be used to scan patient barcodes during diagnostic field studies.

### Optical Resolution

The NTDscope optical system consists of a reversed-lens system [38] with lenses of f-number f/1.8 and a camera sensor with a quad-Bayer array as described above [41, 43]. The green pixels in the array have a peak transmission of ∼525nm, and for that wavelength theoretical on-axis optical resolution is, per the Rayleigh formula, *δ*r^∼^=1.2µm. Actual obtained resolution is limited by undersampling of the image by the sensor pixel array; typical mobile-phone cameras are designed to have significant modulation transfer function (MTF) at Nyquist (i.e., undersampling) to increase field of view at the expense of aliasing. In the case of the NTDscope, for an f/1.8 undersampling is >4.8X in the green, based on the 2.3µm pitch (for green pixels) and the magnification, wavelength, and f-number based Nyquist sampling pitch requirement. This leads to a rough expected (aliasing-limited) on-axis resolution of *δ*r ∼ 5.5µm, consistent with measurements using a custom glass resolution target the size of a capillary, containing numerous arrangements of the 1951 USAF target (Opto-Line International), which yield consistent on-axis resolution better than 4.4µm (228 lp/mm, the limit of the resolution target, Fig 1D). The slightly better-than-expected on-axis resolution is attributable to the uncertainty in estimating sampling-limited resolution as well as the image processing done in the camera firmware based on the additional true 0.8µm pitch of the pixels in the quad-Bayer arrangement. Optical resolution at the extreme edge of the field of view (at 4mm radius) is expected to degrade by a factor of about ⪎ 1.6 (to *δ*r ∼1.8µm along the radial direction) due to NA-limiting from pupil obliquity and increased image distance based on the ∼37° maximum field angle. USAF-target data for extreme field radii are consistent with this, limited again to ≥4.4µm (optically, by expected aliasing issues as discussed, and, in terms of measurement, by the available feature sizes of the test target.) Focus gradients due to imperfect stage alignment in these (small-batch produced, and thus larger-toleranced) devices resulted in worst-case resolution of ∼10µm at the extrema of the FOV (measurements taken at an average 2.7mm from the FOV center, (Fig 1E.iv.), which remains more than sufficient for use with ∼100µm or greater sized filarial worms as well as for *Schistosoma* and soil-transmitted helminth eggs.

### Brightfield and Darkfield Illumination

The system is illuminated by an inner ring consisting of 74 LEDs and an outer ring of 35 LEDs. Illumination of the inner ring is used to produce brightfield imaging, while illumination of the outer ring allows for darkfield capabilities, as shown in Fig 1C. Directly above the illumination LEDs, a diffuser holder hosts a brightfield diffuser, creating uniform illumination across the field of view, and a black, darkfield diffuser, which is used to prevent reflections during darkfield illumination while allowing transmission of brightfield LED illumination. A dome-shaped plastic part, the darkfield dome, surrounds the illumination LEDs and directs light from the outer LED ring towards the edges of the field of view during darkfield illumination.

### Fluorescence Illumination and Emission

Fluorescence illumination uses a laser (405 nm, 50 mW), a mirror, and a set of interference filters suitable for fluorophores such as Alexa Fluor 405, Alexa Fluor 430, ATTO 390, ATTO 425, ATTO 465, etc. The laser is positioned parallel to the NTDscope sample holder, such that the beam hits a mirror oriented at 45° to direct the beam towards the edge of the sample. The excitation filter (CT405/10x, Chroma Technology Corp) is attached directly to the laser. The emission filter (CT450lp, Chroma Technology Corp) is positioned between the reversed lens and the camera module on the collection optics. A diagram showing fluorescence illumination is shown in Fig 1C.

### Sample Capillary and Translation

The NTDscope sample carriage accepts capillaries or other samples with outer dimensions 12mm x 59mm x 1mm, compatible with the original LoaScope capillaries. The carriage moves in one dimension and is able to capture seven distinct FOVs. The original capillary, developed for imaging *Loa loa* microfilariae (mf) in peripheral blood, has a channel with dimensions of 54mm x 4mm x 200µm (Fig 1A), holding a total blood volume of 43.2µL. Each of the seven distinct FOVs that can be imaged covers a volume of 4.96µL of blood, for a total of 39µL per capillary. In addition to the rectangular capillary, we iterated on the design to create capillaries for other use cases, conserving external dimensions to retain compatibility with the NTDscope carriage. For example, a tapered capillary for processing urine and stool samples, shown in Fig 3, and a thick smear capillary, shown in Fig 4.

### Software

The NTDScope runs on Android 9 operating system (OS). Given the operating system, machine learning (ML) algorithms tailored to specific neglected tropical diseases can be loaded onto the device for automatic pathogen detection [44, 45]. We developed custom Android applications (apps) for the NTDscope, enabling control of the sample location, focus, capture of images and videos, etc. The location of the sample is controlled via software by rotating a servomotor gear attached to the capillary carriage. Focus is controlled by vertical translation of the camera lens module. Brightfield and darkfield illumination are controlled via software by turning on the central (brightfield) or outer ring (darkfield) LEDs. Flourescence illumination is achieved by activating the laser. A custom app was developed for the quantification of *Loa loa* burden, and was tested in rural field settings in Cameroon and Gabon. This app allows the user to capture patient information (with an option to read patient-associated barcodes), and insert a blood-filled capillary, after which it automatically captures five-second videos in seven different locations along the capillary, translating it before each new acquisition. After imaging, the sample videos are processed and the *Loa loa* burden is quantified and reported.

## Results

### Quantification of live microfilariae in blood samples

The NTDscope can be used to capture videos of moving samples, such as microfilariae (mf) of *Loa loa* and other filarial parasites. The videos can be processed to detect live parasites without the need for staining or additional sample preparation, allowing for point-of-care diagnosis and quick time-to-result. We demonstrate this by using the original rectangular capillaries to acquire videos of microfilariae in blood and plasma, as shown in Fig 2.

**Fig 2.**
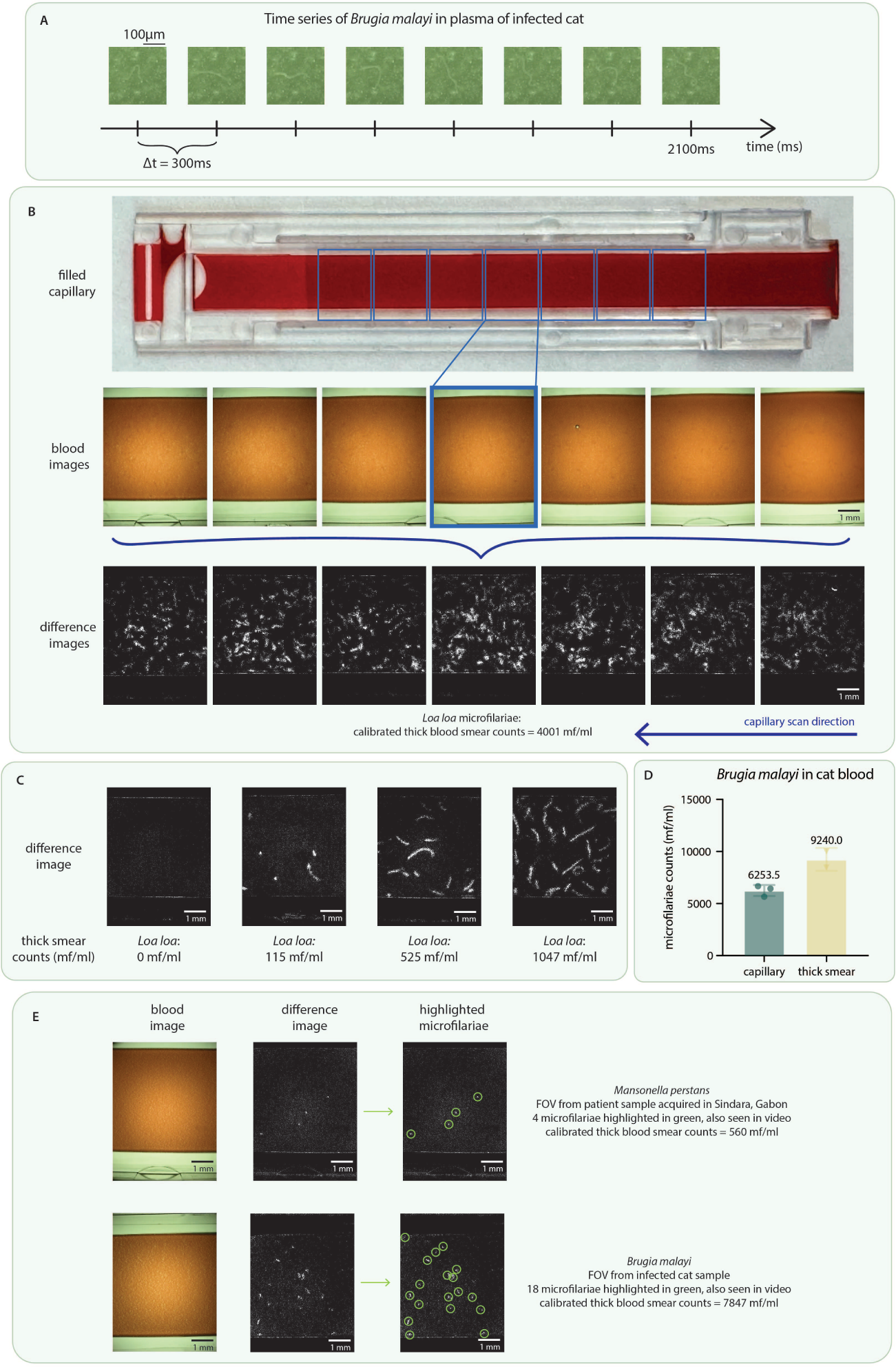
NTDscope for live imaging of microfilariae in blood and plasma samples. **A** Time series showing the movement of a *Brugia malayi* microfilaria in plasma of an infected cat. **B** (top) image of a capillary filled with peripheral blood of a human patient infected with *Loa loa*, collected and imaged in Cameroon, (middle) screenshots of the videos corresponding to the 7 FOVs imaged for the capillary, (bottom) difference images generated from the 7 videos. The patient had calibrated thick smear counts of 4001 mf/ml. A video of the middle (highlighted) FOV is shown in Supplementary Video 1.**C** Difference images generated from videos from four different patients in Cameroon, three of which were infected with *Loa loa* mf (thick smear counts 115-1047 mf/ml) and one patient who was negative for *Loa loa*. **D** Comparison of microfilariae counts from a calibrated thick smear and counted manually from videos acquired on the NTDscope, for a blood sample infected with *B. malayi* mf. **E** Snapshots of videos and corresponding difference images of samples containing *M. perstans* (top) and *B. malayi* (bottom) mf. The mf identified in videos and difference images are circled in green. Data was processed and visualized using GraphPad Prism 10.

We used samples of patients infected with *Loa loa* and *Mansonella perstans*, as validated by calibrated thick blood smears, acquired in Cameroon and Gabon. To fill the capillaries, a patient fingertip was pricked with a lancet and peripheral blood was wicked into the capillaries via capillary action (as described in [36]). The blood-filled capillaries were then inserted into the NTDscope, and five-second videos of seven distinct FOVs of each sample were captured, with the capillary automatically translated by the servomotor between each imaged FOV. Videos are automatically analyzed for mf detection using software loaded on the device. The total imaging and processing time for diagnosis is less than 3 minutes. Fig 2B shows a picture of one capillary filled with blood from a *Loa loa*-positive patient in Cameroon, followed below by screenshots of the 7 videos acquired (blood images), and showing the capillary regions that the FOVs correspond to. A video of one of these FOVs is shown in Supplementary Video 1.

**Fig 3.**
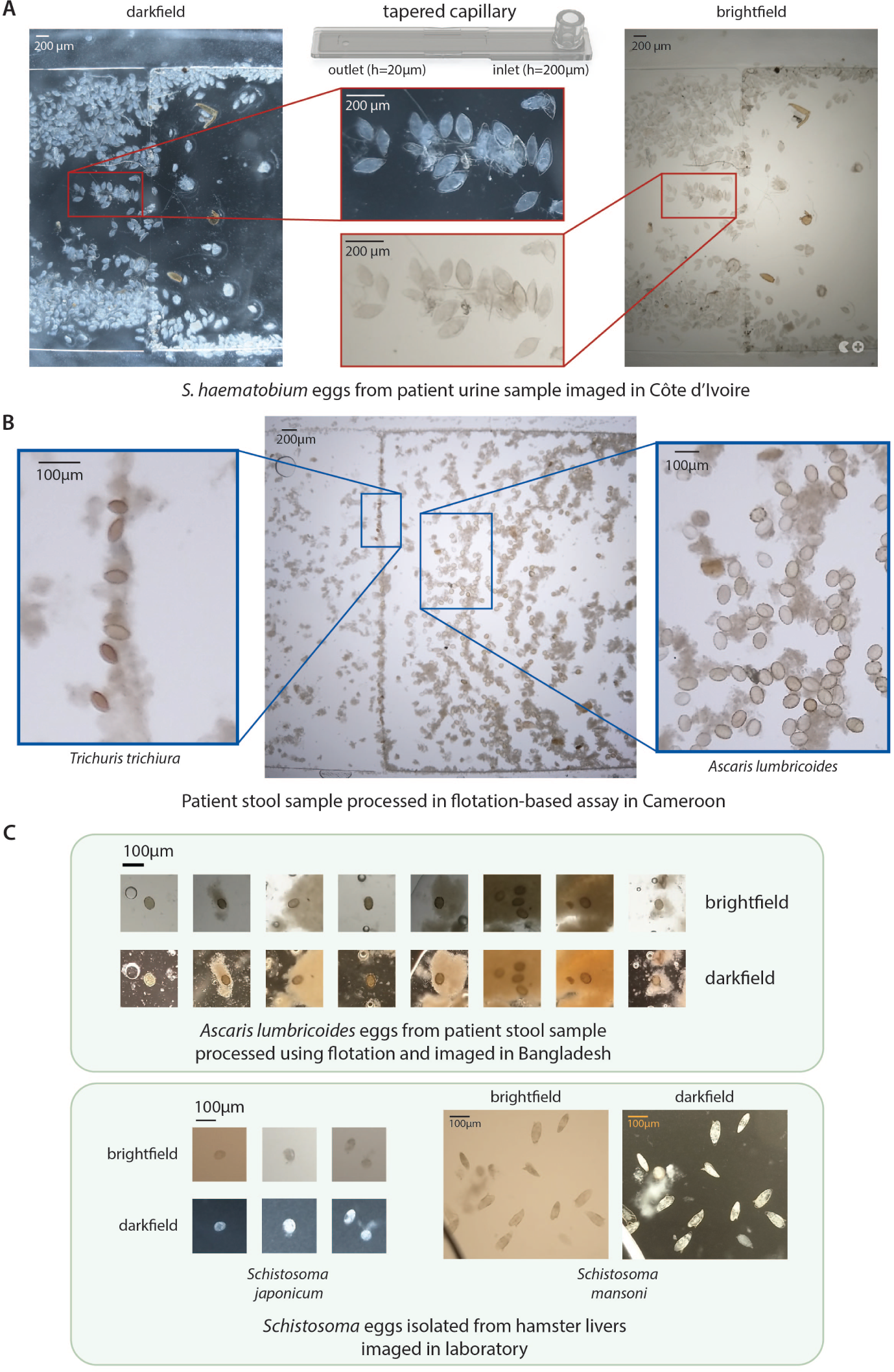
NTDscope can image parasitic eggs in urine and stool. **A** Images of *Schistosoma haematobium* eggs from a urine sample of a patient in Côte d’Ivoire, acquired using brightfield (BF) and darkfield (DF) imaging. **B** Image of soil-transmitted helminth (STH) eggs from a patient stool sample processed using a flotation-based assay and imaged using the NTDscope in Cameroon. Insets showing *Trichuris trichiura* (left) and *Ascaris lumbricoides* (right) eggs captured in different locations of the NTDscope tapered capillary. **C** (top) images of *A. lumbricoides* eggs from a patient stool sample, processed with a flotation solution and imaged using BF and DF illumination on the NTDscope in Bangladesh. (bottom) *Schistosoma japonicum* and *Schistosoma mansoni* eggs isolated from hamster livers and imaged on the NTDscope using BF and DF illumination.

Patient sample videos were processed by an algorithm that subtracts subsequent frames to generate difference images (as described in [36]). Difference images show regions of high intensity in locations of the FOV where there was movement, in this case due to the presence of microfilariae in the sample. The bottom row of Fig 2B shows the difference images generated from the videos of the patient sample, which had calibrated thick smear counts of 4001 mf/ml. The higher number of high intensity regions in difference images corresponds to higher *Loa loa* mf concentration in patient samples. This is demonstrated in Fig 2C, where the difference image corresponding to one FOV of four different patients is shown. The corresponding thick smear counts for those patients (ranging from 0-1047 mf/ml) are shown below each image.

We also used blood and plasma samples of cats infected with live *Brugia malayi* microfilariae (provided by the Filariasis Research Reagent Resource Center for distribution through BEI Resources, NIAID, NIH). For these samples, blood and plasma were pipetted directly into the NTDscope capillaries. To capture videos of plasma, the cat blood was left to separate into its components overnight, after which 43.2 µL of the supernatant (corresponding to plasma) were pipetted into a NTDscope rectangular capillary for video acquisition. Snapshots of this video are shown in Fig 2A, and the full video can be found in Supplementary Video 2.

To determine the expected correlation between actual microfilariae concentration in a sample and the number of microfilariae that could be identified from the videos acquired on the NTDscope, we prepared three capillaries and two calibrated thick smears from a sample of a cat infected with *B. malayi*. We then manually counted worm movements in the videos and counted the visible mf in the calibrated thick smears. The thick smear and manual capillary counts are shown in Fig 2D. There is a decrease in the manual capillary counts compared to the thick smears, which was also seen in [36], and could be due to a fraction of dead mf that are counted in the thick smears but are not captured in difference images, or due to live mf that do not make it into the capillaries due to their shape and size.

Different filarial species have distinct movement signatures, which can be seen by examining difference images from samples with mf of different species. Fig 2E shows the difference images of a sample of a human patient infected with *M. perstans*, collected in Gabon, and difference images of a blood sample of a cat infected with *B. malayi*. The intensity peaks corresponding to the location of mf movement in the FOVs are marked by green circles. The peaks are smaller for *M. perstans* when compared to those produced by *B. malayi* and even *Loa loa* (seen in Fig 2B and C). This implies that *M. perstans* microfilariae consistently create much smaller areas of blood displacement as compared to *Loa loa* and *B. malayi*. These signatures could eventually be used to distinguish between filarial species.

### Multi-contrast imaging of helminth eggs

The NTDscope can be used to image parasite eggs in urine and stool samples with minimal processing. For imaging of *Schistosoma haematobium* eggs in patient urine samples, we used a tapered capillary described in [32] and shown in Fig 3A. The capillary has a rectangular channel with a height that tapers down from 200µm near the inlet to 20µm near the outlet hole. These capillaries can be connected to a syringe to easily flow liquids through, including urine and stool processed with a flotation solution. The tapered design ensures that parasite eggs are trapped along the channel, at a location dependent on their size. The external dimensions of the tapered capillary are such that it can be inserted into the NTDscope carriage for imaging. Fig 3A shows *S. haematobium* eggs imaged using brightfield and darkfield illumination on the NTDscope. The eggs are from patient urine samples collected in Côte d’Ivoire.

The NTDscope can also be used to image soil-transmitted helminth (STH) eggs found in patient stool samples. STH is traditionally detected using the Kato-Katz method, a time-consuming technique that involves sieving, staining and smearing a stool sample onto a glass slide for imaging using a microscope [46]. An improvement on this method has been the Mini-FLOTAC technique [47], in which the stool is homogenized and mixed with a flotation solution (using a kit called the Fill-FLOTAC), such that eggs can be concentrated and separated from stool particulates. After flotation, the samples are loaded into the Mini-FLOTAC apparatus, which is used for microscopic examination of the sample and identification of parasitic eggs. We used the FLOTAC technique to process stool samples taken from a patient with known STH infection in Cameroon. The sample was homogenized and mixed with a flotation solution using a Fill-FLOTAC. Then, instead of loading the sample into a Mini-FLOTAC and imaging on a microscope, we injected the floated stool solution into the NTDscope tapered capillary. This capillary is particularly useful for samples that contain multiple STH species, which are separated into different locations of the capillary due to their different sizes. As seen on the image in Fig 3B, the bigger *Ascaris lumbricoides* eggs (45-75 µm thick) were concentrated on the right side of the image, corresponding to the region of the capillary closest to the inlet, while the smaller *Trichuris trichiura* eggs (20-25 µm thick) were concentrated on the left side, closest to the capillary outlet hole.

We also explored an alternative, simpler, method for stool sample processing using the Fecalyzer, a device that is similar to the Fill-FLOTAC but is smaller, cheaper and more commonly used for veterinary applications [48]. To prevent clogging issues on the tapered capillary due to unfiltered stool particulates, we developed a prototype capillary using two thin pieces of laser-cut acrylic coated with a hydrophilic solution (Tetronic 904 dissolved in isopropanol alcohol), that could be inserted into the NTDscope for imaging. We used the Fecalyzer to homogenize 0.2 grams of stool and added ∼14 mL of a zinc sulfate (ZnSO4) flotation solution such that a meniscus formed. We placed the coated acrylic pieces on top of the meniscus and waited 15 minutes to allow the STH eggs in the stool sample to float to the top and adhere to the acrylic surface. The acrylic pieces were then removed and inserted into the NTDscope for imaging. Fig 3C (top) shows *A. lumbricoides* eggs from patient stool samples collected and processed as described above, in an STH-endemic area in Bangladesh. In addition to decreasing cost, sample preparation steps, and required materials for STH detection, this is an example of how new assays and sample processing techniques can be easily adapted for imaging on the NTDscope.

Lastly, we show that the NTDscope can be used to image *Schistosoma* eggs of different species, including *S. japonicum* and *S. mansoni*. We used viable schistosoma eggs extracted from livers of infected hamsters (provided by the Schistosomiasis Resource Center of the Biomedical Research Institute (Rockville, MD)) and observed that the NTDscope resolution is sufficient for the eggs of different species can be differentiated by eye on the NTDscope images (as shown in Fig 3C (bottom). We also acquired videos of *S. mansoni* miracidia hatched from liver-extracted eggs in brightfield and darkfield illumination, one of which is shown in Supplementary Video 3.

### Imaging of stained patient samples

We explored the use of the NTDscope for imaging of fixed and stained slides, which could be useful in clinical laboratory settings where fixing and staining of samples is routine. We first developed an NTDscope-compatible capillary to simulate a thick smear (Fig 4A). This capillary is made from laser cut-pieces of acrylic and is coated with a hydrophilic solution (Tetronic 904 in isopropanol alcohol). The capillary holds a fixed volume of blood (30µL) and can be fixed and stained following a standard Giemsa staining protocol. We prepared a thick smear capillary using a blood sample of a cat infected with *B. malayi* microfilariae, stained it using Giemsa and imaged it with the NTDscope. Given the large field of view of the NTDscope, only seven images were required to cover the entire sample. The stiched images are shown in Fig 4A. The NTDscope could resolve the *B. malayi* microfilariae well, and this is another demonstration of how the NTDscope can be adapted for other relevant assays.

**Fig 4.**
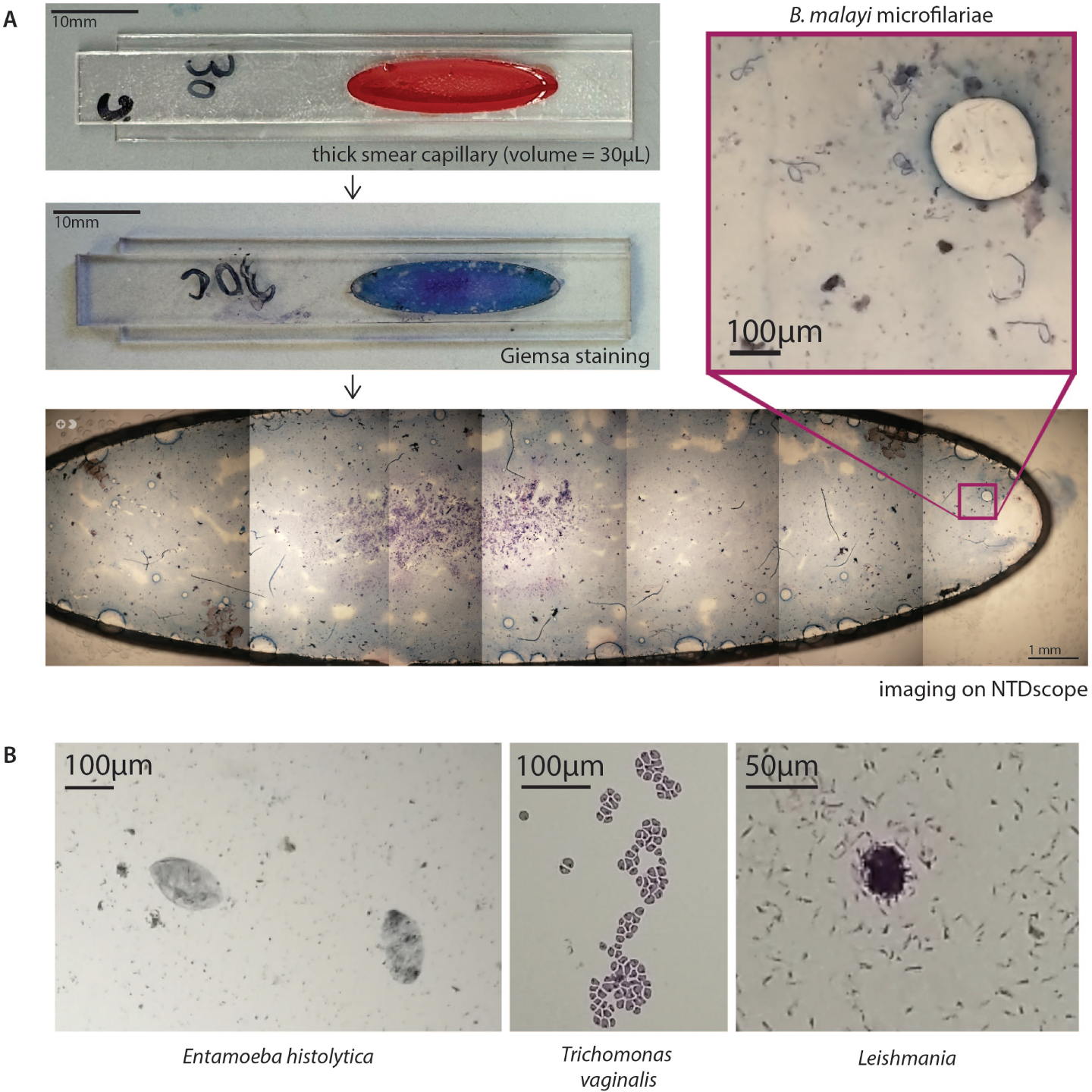
The NTDscope can image parasites on fixed and stained slides. **A** (top left) picture of a thick smear capillary designed to be imaged with the NTDscope, including 30µL of blood from a cat infected with *Brugia malayi*. (middle left) Thick smear capillary after Giemsa staining. (bottom) stitched images of Giemsa-stained capillary, collected with the NTDscope, with an inset showing 6 *B. malayi* microfilariae (top right). The full video shown in Supplementary Video 3. **B** Images of fixed and stained slides with parasites, collected on the NTDscope, showing (left) *Entamoeba histolytica*, (middle) *Trichomonas vaginalis*, and (right) *Leishmania donovani* promastigotes.

To explore whether the NTDscope could be used for imaging other relevant parasites, we modified a device slightly such that it could fit standard microscope slides and imaged fixed and stained slides sourced from Carolina Biological. We imaged *Entamoeba histolytica*, *Trichomonas vaginalis*, and *Leishmania donovani* promastigotes. The *L. donovani* promastigotes were too small (1-2µm) to fully resolve with the NTDscope; computational techniques, such as dithering, could improve the resolution of the NTDscope such that these and other parasites (e.g. malaria) can also be imaged on the device [49].

### Imaging of lateral flow assays

To test the ability of the NTDscope to quantify lateral flow assays (LFAs), we imaged two commercially-available lateral flow assay strips for severe acute respiratory syndrome coronavirus 2 (SARS-CoV-2) antigen detection, one processed using a positive patient sample and one using a negative patient sample. The lateral flow strips were removed from their plastic housing and attached to a glass support with dimensions of the NTDscope carriage. The patient nasal swab collection and processing was done according to the manufacturer, and the three drops of swab solution and assay components were added to the strips and immediately imaged using the NTDscope. For the positive sample, images of the test line were acquired every minute for five minutes. End-point images were also acquired 15 minutes after the start of the assay.

Figure 5A.i. includes a time-lapse of the test band for the positive patient, showing how the intensity of the band increases in time (as shown by a decrease in pixel intensity, Fig 5A.ii.). We also show the control and test bands for two positive samples and one negative sample (Fig 5A iii.) and a scan of the entire strip showing the control and test band for a positive patient.

**Fig 5.**
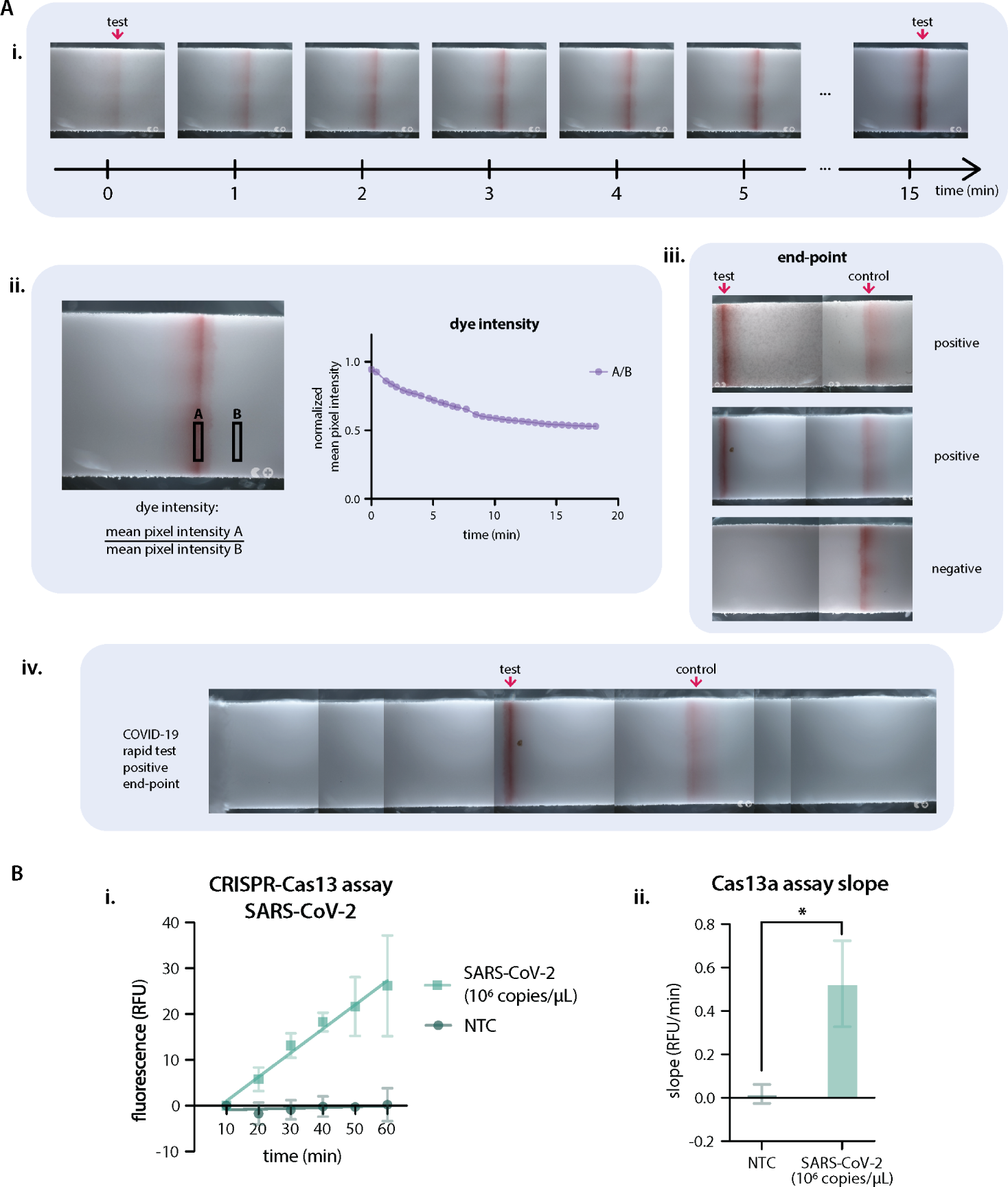
The NTDscope can be used to image lateral flow test strips and CRISPR-Cas molecular assays. **A** Lateral flow assays imaged on the NTDscope. A.i. Time series (15 min) of the test line on a COVID-19 antigen test, tested on a positive patient. A.ii. Measurement of the dye intensity over time, of the test line shown in A.i., calculated by dividing the mean pixel intensity of a rectangular region of interest (ROI) over the test line by an ROI outside of the test line. Pixel intensity was measured using Fiji (ImageJ). A.iii. End-point images after 15 minutes for three COVID-19 tests (2 positive, 1 negative), showing the test and control lines. A.iv. Scan of the entire COVID-19 test strip, showing the control and test bands for a positive patient after 15 minutes. **B** Results of a CRISPR-Cas13 assay run on NTDscope. B.i. Measurement of fluorescence intensity over time for a Cas13 assay with a guide RNA specific to SARS-CoV-2, in the presence (SARS-CoV-2 1e6 copies/µL) and absence (non-target control — NTC) of synthetic SARS-CoV-2 RNA. Fluorescence intensity was measured during the first 60 minutes of assay time, measurements were acquired every 10 minutes. The data from the first 10 minutes was discarded to allow assay components to equilibrate to temperature. Experiment performed in triplicate. B.ii. Slopes of the curves calculated by performing simple linear regression of the data for each replicate of B.i., slopes of the positive samples were compared to the no target RNA controls (NTC) using an unpaired t-test. * is p*≤*0.05. Data were processed and visualized using GraphPad Prism 10.

### Imaging a CRISPR-Cas13a SARS-CoV-2 assay

Detection of RNA or DNA with Clustered Regularly Interspaced Short Palindromic Repeats (CRISPR) is a promising approach for diagnosis of infectious diseases [50]. To demonstrate ability of the NTDscope to detect a CRISPR reaction, we leveraged the device’s fluorescence capabilities to image a CRISPR-Cas13a assay for the detection of SARS-CoV-2 (Fig 5B).

As previously described [51], we used CRISPR-Cas13a with a CRISPR RNA (crRNA, also known as a guide RNA) to recognize a specific sequence in the SARS-CoV-2 N gene. Briefly, the crRNA is incubated with the Cas13a enzyme to form a ribonucleoprotein that is activated to nonspecifically cleave a ssRNA reporter-quencher upon target sequence recognition. The enzyme Cas13a was expressed and purified as previously described, and the crRNA for SARS-CoV-2 and a synthetic target were ordered from Synthego [51]. The ssRNA reporter-quencher was modified from [51] to include a 5’ ATTO425 fluorophore and a 3’ IowaBlack FQ quencher. The assay was prepared by preassembling Cas13a-crRNA RNP complexes for 15 minutes at room temperature, after which the ssRNA reporter and the target RNA at either 1 × 10^6^ copies of RNA per µL (cp/µL) for positive controls or 0 cp/µL for the non-target control (NTC) were added.

For NTDscope fluorescence measurements, the CRISPR-Cas13a reactions were loaded individually into two-5µL glass capillaries (Vitrocom, cat# 5010-050), which were attached to a modified glass slide (12mm x 75mm x 1mm) using a double-sided adhesive (ARcare 90445Q) to fit the NTDscope carriage. In each of the three glass slides prepared, there were two glass capillaries, containing two reactions (Positive and NTC). We used the fluorescence imaging capabilities of the NTDscope to image each glass slide every 10 minutes for 60 minutes after equilibration to the assay temperature of 37°C. Fluorescence signal was quantified using ImageJ Fiji by taking an average of pixel intensity of a fixed pixel area across all images in the time series for the positive and the NTC reactions. A plot of the fluorescent signal over time was generated using the measured pixel intensities (Fig 5B.i.). Slopes of the curves were calculated by performing simple linear regression of the data for each replicate (Fig 5B.ii.). Slopes of the positive samples were compared to the NTC using an unpaired t-test.

The slope of the fluorescence increase for the positive sample measured on the NTDscope was significantly larger than the slope of the non-target control, confirming SARS-CoV-2 RNA detection by Cas13a. Additionally, the slope of the line for 1 × 10^6^ cp/µL produces similar results to slopes seen when the assay was run on a temperature-controlled plate reader (Supplementary Fig 1). To make the plate reader measurements, 15 µL reactions were loaded into individual wells from a 384-well plate and incubated in a plate reader (TECAN, Spark) for 60 minutes at 37°C, with fluorescence intensity measurements taken every 2.5 minutes (*λ_ex_*: 425 nm; *λ_em_*: 500 nm). The slope of the reactions starting at 10 minutes (to account for assay temperature equilibration) was calculated by linear regression of the mean background-subtracted reaction rate (RFU/sec) with SEM. The plate reader results are shown in Supplementary Fig 1. All data were compared using an unpaired t-test.

## Discussion

In this work, we presented the design of the NTDscope, a portable, multi-contrast microscope that enables diagnosis of neglected tropical diseases at the point-of-care. We demonstrated its imaging capabilities for microfilariae in peripheral blood, *Schistosoma* eggs in urine samples, and STH eggs in processed stool samples. We also showed that the device is capable of reading molecular assays by imaging both lateral flow assays and fluorescence-based CRISPR-Cas assays. Sample preparation is simplified for these applications by the use of disposable plastic capillaries, including rectangular straight channel capillaries for defined blood volumes, tapered capillaries for filtering eggs from urine and floated stool samples, and as well as a prototype capillary for thick blood smears that is compatible with the NTDscope.

As new diagnostic applications are developed for the NTDscope, new capillary designs that fit the carriage dimension and focal plane can be designed. The original straight channel capillary was designed to easily load a peripheral blood sample for quantification of microfilariae, but it can also be loaded with other liquids for other uses. Previous applications have included imaging of parasitic eggs isolated from a flotation solution using the Fill-FLOTAC or Fecalyzer devices, as well as imaging of diatoms in marine samples for educational purposes. The tapered capillary that was designed to isolate *Schistosoma* eggs from urine has also proven useful for other applications that require size-exclusive filtering and concentration, such as separating two different species of STH eggs.

The prototype thick smear capillary demonstrates potential for integrating common clinical laboratory assays into the device, including wet mounts for protozoa and fungi, three-part blood counts, and parasitic egg viability assays [52, 53]. The ability to capture images of live parasites has the potential to provide new information about infections and response to treatment. For example, NTDscope videos of *Loa loa* microfilariae motion inherently capture only live parasites rather than dead microfilariae that could appear in conventional thick smear microscopy. In the case of low parasitemia, additional FOVs and multiple capillaries from a single patient can be analyzed, which could improve sensitivity for treatment stopping decisions in near-elimination settings.

The diagnostic power of the NTDscope is enhanced by integrating machine learning (ML) for automated analysis of patient samples. Importantly, use of ML can significantly reduce the time required for sample analysis, including manual counting of parasites that can take a minimum of 10 minutes per slide for well-seasoned technicians [54] and can have high variability [55, 56], reducing fatigue as well as increasing throughput. Use of the Android OS environment allows existing ML models designed for mobile applications to be easily run on the device. Furthermore, the portability of the NTDscope simplifies data collection for NTDs, increasing the amount of available training data for ML models, which is particularly important for capturing patient samples in NTD-endemic regions where microscopes with digital cameras are unavailable. Beyond classification of parasites in static images, ML models have the potential to differentiate microfilarial species based on movement signatures, which we have observed to be different for *M. perstans*, *B. malayi*, and *Loa loa* microfilariae (Fig 2). As more field data is collected, ML algorithms could be developed to distinguish between multiple filarial species. For some diagnostic uses cases, visualizing an ML-highlighted parasite on the screen of the NTDscope could help to build confidence in the tool for both providers and patients.

The multi-contrast imaging capabilities of the NTDScope could prove useful for future diagnostic applications that involve species differentiation, such as for distinguishing soil transmitted helminth species and *S. mansoni* eggs in stool samples. Darkfield imaging has proved beneficial when used for ML-based identification of *S. haematobium* eggs, where models trained on darkfield images performed better than those trained on brightfield images [34]. Darkfield illumination is particularly helpful when distinguishing eggs from other debris found in samples, which is typically achieved through staining, requiring extra steps and assay components. Given that many parasitic eggs and worms exhibit autofluorescence [57–60], the NTDscope could further facilitate speciation with its fluorescence capabilities.

The fluorescence capabilities of the NTDscope provide the potential to replace bulky laboratory equipment, such as plate readers for molecular assays. Fluorescence-based molecular assays, such as those based on CRISPR-Cas enzymes, are of growing interest for infectious disease diagnosis as they avoid the need for conventional PCR machines and temperature cycling (amplification-free CRISPR-Cas reactions are isothermal). CRISPR-Cas diagnostic assays have been demonstrated for the detection of neglected diseases including Schistosomiasis [61, 62], Trypanosomiasis [63], Leishmaniasis [64], Malaria, [65], Zika, Dengue [66], and others [67–69]. These reactions can be read using portable devices [51, 70], and could be adapted to be read using the NTDscope. We also showed that the NTDscope can be used to read the results of lateral flow assays. While multiple portable LFA readers exist [71–76], the potential to read test strips with a magnified imaging system could improve sensitivity in cases of light infection, help digitize results for future use, and open the possibility of combining multiple tests onto a single test strip, reducing the quantity of reagents needed [77–82].

Presently, the main limitation of the NTDscope lies in device availability. The NTDscope presented here is a prototype built to demonstrate the feasibility of a fully-integrated diagnostic device for multiple NTDs, and 80 devices were built in a limited manufacturing run. Collaborators in Cameroon, Gabon, Côte d’Ivoire, and Bangladesh continue to work with the device to establish efficient workflows for expanded use, as well as explore additional diagnostic and disease mapping applications. With additional feedback from ongoing field work, future versions of the device have the potential to be manufactured or assembled in the countries where they are used. The NTDscope represents an important step toward increasing access to disease testing in regions with limited healthcare infrastructure through portable diagnostic technology, a direction that may also benefit regions beyond those with NTDs.

## Conclusion

The NTDscope is a portable, multi-contrast microscope designed to diagnose neglected tropical diseases at the point-of-care. It features high-resolution imaging at a low cost, achieved by taking advantage of high-quality, mass-produced consumer electronic parts, such as mobile phone lenses. The device is compact, user-friendly, and doesn’t require mains electricity to run. Here, we demonstrate use of the NTDscope for a diverse set of assays and samples in laboratory and field settings including Cameroon, Gabon, Bangladesh, and Côte d’Ivoire. When combined with machine learning for automated diagnostics, the NTDscope has the potential to increase the reach and impact of healthcare professionals. The portability of the NTDscope can also increase the amount of image data collected from patients with NTDs, a necessary step for training of machine learning models that has been difficult to achieve in NTD-endemic regions. We envision the NTDscope as a platform technology that addresses diagnostic needs for multiple NTDs and supports ambitious disease control and elimination programs.

## Data Availability

Data for this manuscript can be accessed in the following location:
https://drive.google.com/drive/folders/11_Lq0iQNtYUVxrnlKnjKTdogHz93VEcn?usp=sharing

## Supporting information

**Supplementary Figure 1. CRISPR-Cas13a assay results on plate reader**. i. Measurement of fluorescence intensity over time for a Cas13 assay with a guide RNA specific to SARS-CoV-2, in the presence (SARS-CoV-2 1e6 copies/µL) and absence (non-target control — NTC) of synthetic SARS-CoV-2 RNA. Fluorescence intensity was measured on a Tecan Spark plate reader during the first 60 minutes of assay time, measurements were acquired every 10 minutes. The data from the first 10 minutes was discarded to allow assay components to equilibrate to temperature. Experiment performed in triplicate. ii. Slopes of the curves calculated by performing simple linear regression of the data for each replicate of i., slopes of the positive samples were compared to the no target RNA controls (NTC) using an unpaired t-test. Data are expressed as mean +-standard error of the mean (SE). * is p<0.05; ** is p<0.01; *** is p<0.001; **** is p<0.0001. Data was processed and visualized using GraphPad Prism 10.

## Acknowledgments

We thank all of our partners and field collaborators for making this work possible. We thank the researchers and clinical staff at CERMEL, ISM, the Centre Suisse de

---

**Supplementary Video 1. *Loa loa* microfilariae in peripheral blood.**

Video showing a field-of-view (FOV) of a capillary filled with peripheral blood of a human patient infected with *Loa loa*, collected and imaged in Cameroon. This corresponds to the fourth FOV from left to right in Fig 2B. The patient had calibrated thick smear counts of 4001 mf/ml.

**Supplementary Video 2. *Brugia malayi* microfilariae in plasma.**

Video showing multiple *B. malayi* microfilariae in plasma of an infected cat.

**Supplementary Video 3. *S. mansoni* miracidia**

Video showing *S. mansoni* miracidia hatched from eggs extracted from livers of infected hamsters. The sample was imaged under brightfield and darkfield illumination.

Recherches Scientifiques en Côte d’Ivoire, and the Centre for Diarrhoeal Disease Research, Bangladesh (icddr,b), for their invaluable help and support. We would also like to thank all of the patients who provided us with samples.

The *Brugia malayi* in cat blood was provided by the NIH/NIAID Filariasis Research Reagent Resource Center for distribution through BEI Resources, NIAID, NIH: Brugia malayi Microfilariae in Cat Blood, Live, NR-48887. The *Schistosoma* eggs were provided by the Schistosomiasis Resource Center of the Biomedical Research Institute (Rockville, MD) through NIH-NIAID Contract HHSN272201700014I. We thank Sarah Schmid from the Schistosomiasis Resource Center for her experimental help.

## Author contributions

- Conceptualization: MDDLD, ZLM, NAS, DHF, MVDA, DAF
- Data curation: MDDLD, ZLM, CFN, AMB, ED, DC, HCND, LDY, VP, RZM, JTC
- Data analysis: MDDLD, ZLM, CFN, AMB, NAS
- Funding acquisition: MDK, ALLN, MLO, JK, MR, IIB, JTC, DHF, MVDA, DAF
- NTDscope design: NAS, DHF, MVDA, DAF
- NTDscope assembly: MDDLD, CFN, JPC, MDK, ALLN, NAS, DHF, MVDA, DAF
- Software: DB, CBD, JGV, ED, DBP, DC, MLO, MVDA
- Supervision: MDK, ALLN, HCND, JK, RZM, MR, IIB, JTC, NAS, DHF, MVDA, DAF
- Field validation: MDDLD, ZLM, AMB, MDK, ALLN, JGV, ED, DBP, DC, MLO, HCND, JK, LDY, VP, SDD, RZM, IIB, JTC, AK, MK, ZN, RH
- Writing – original draft: MDDLD, ZLM, CFN, NAS, DAF
- Writing – review & editing: all authors

## Notes

### Competing Interest Statement

The authors have declared no competing interest.

### Funding Statement

DAF received funding from the Gates Foundation (INV-008782). DAF received funding from the Chan Zuckerberg Biohub, Wagner Foundation and the Igarashi Research Fund. The sponsors did not play any role in study design, data collection and analysis, decision to publish, or preparation of the manuscript.

### Author Declarations

This work contains images and videos of patient samples from four separate studies conducted in Cameroon, Gabon, C’te d'Ivoire, and Bangladesh. The study in Cameroon was conducted in the Awae Health District, located in the Mefouet-Afamba Division, between September and October of 2023. Finger prick blood was collected from study participants. Ethical permission for this study was granted by the Centre Regional Ethical Committee for Research on Human Health (CRERSH-Ce; CE No0094/CRERSHC/2023) and administrative authorization was obtained from the Centre Regional Delegation for Public Health of the Ministry of Public Health. The study participants signed informed consents. The study in Gabon was conducted in the Sindara and Lambaréné regions in August of 2023. Finger prick blood was collected from study participants. Ethical permission for this study was granted by the Comité d'ethique Institutionnel du CERMEL (CEI-026/2022). Adults 18 years or older were invited to participate and provided a written informed consent. The study in C’te d'Ivoire was conducted around the town of Azaguié in January of 2024. Ethical permission was granted by the University Health Network, Toronto, Canada (REB #21-5582) and the Comité National d'éthique des Sciences de la Vie et de la Santé, Abidjan, C’te d'Ivoire (REB #186-21/MSHPCMU/CNESVS-km). Ethical permission was also granted by the local health district officer. Community members aged five and older were asked to provide a urine sample. Adults provided written consent and assenting children who had written consent from a parent or guardian were included. The study in Bangladesh was conducted from October 2019 - December 2020 in the Rohingya Refugees camp in Bangladesh. Ethical permission for this study was granted by the International Centre for Diarrhoeal Disease Research, Bangladesh (icddr,b) Institutional Review Board (research protocol #PR-19014). Children who were residents of the camp and seeking care from icddr,b-operated Diarrhoea Treatment Centre were asked to participate by providing stool samples. Children were included if they assented and had written consent from a parent or guardian. Banked stool samples were used for the purposes of this paper; participants and their guardians were informed that unused samples would be stored for future use, and at any time participants and their guardians could contact the study team if they wished for the samples to be discarded and not saved.

