## Supplementary figures and images for "NTDscope: A multi-contrast portable microscope for disease diagnosis"

### Supplementary Figure 1

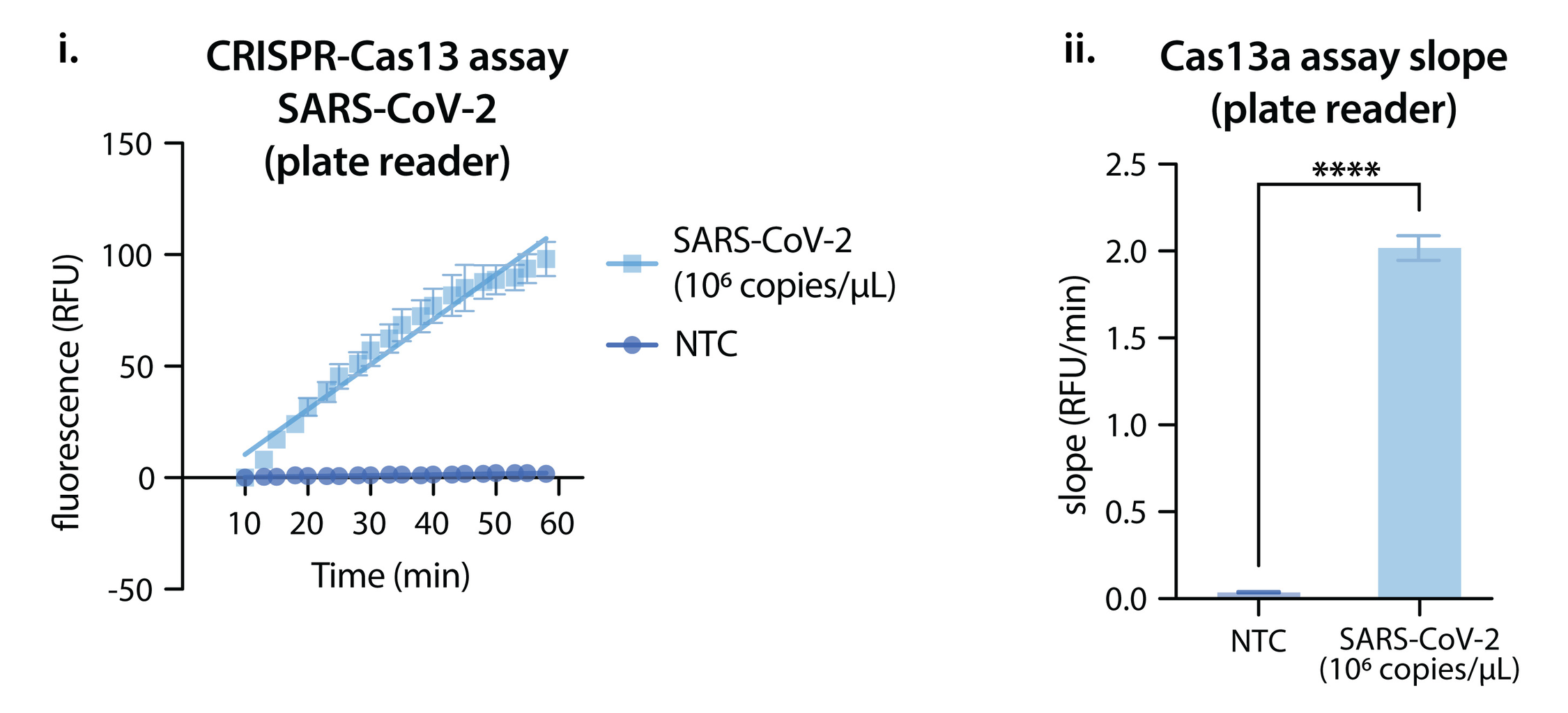
